# *Mycoplasma genitalium* infection and adverse pregnancy outcomes among pregnant women in South Africa: prospective cohort study

**DOI:** 10.64898/2026.08.09.26360025

**Authors:** Ranjana MS Gigi, Mandisa M Mdingi, Hyunsul Jung, Lydia Braunack-Mayer, Eric Mensah, Jean-Benoît Rossel, Chibuzor M Babalola, Christina A Muzny, Christopher M Taylor, Andrew Medina-Marino, Jeffrey D Klausner, Janneke HHM van de Wijgert, Remco P H Peters, Nicola Low

**Affiliations:** Institute of Social and Preventive Medicine, University of Bern, Bern, Switzerland; Research Unit, Foundation for Professional Development, East London, South Africa; Department of Medical Microbiology, University of Pretoria, Pretoria, South Africa; Department of Clinical Research, University of Bern, Bern, Switzerland; Department of Population and Public Health Sciences, Keck School of Medicine, University of Southern California, Los Angeles, California, USA; Division of Infectious Diseases, University of Alabama at Birmingham, Birmingham, Alabama, USA; Department of Microbiology, Immunology, and Parasitology, Louisiana State University Health Sciences Center, New Orleans, Louisiana, USA; Department of Psychiatry, Perelman School of Medicine, University of Pennsylvania, Philadelphia, Pennsylvania, USA; Julius Center for Health Sciences and Primary Care, University Medical Center Utrecht, Utrecht University, Utrecht, The Netherlands; Division of Medical Microbiology, University of Cape Town, Cape Town, South Africa

## Abstract

**Background:** Sexually transmitted infections (STIs) and vaginal dysbiosis during pregnancy are associated with adverse pregnancy outcomes. *Mycoplasma genitalium* is the most recent STI implicated but evidence remains limited. The objectives of this study were to investigate 1) the association between *M. genitalium* infection during pregnancy and gestational age at delivery, preterm birth, miscarriage or stillbirth, and low birth weight and 2) the interaction with vaginal dysbiosis.

**Methods:** We conducted a prospective cohort study in East London, South Africa. We enrolled pregnant women at gestational age <27 weeks, confirmed by ultrasound. We tested vaginal samples using nucleic acid amplification tests for *M. genitalium*, *Chlamydia trachomatis*, *Neisseria gonorrhoeae*, *Trichomonas vaginalis*, other genital mycoplasmas and *Candida* spp. We defined vaginal dysbiosis using Gram-stain criteria as a Nugent score 4-10. We used quantile regression to compare the outcome in women with and without *M. genitalium* across the gestational age distribution, adjusting for prespecified sociodemographic and clinical characteristics and co-occurring organisms.

**Results:** From April 1, 2021 to August 29, 2023, we enrolled 603 women, followed up 584 and obtained pregnancy outcomes for 560 (93%). Median age was 28 years (interquartile range, IQR 24, 33) and 27% of women were living with HIV. *M. genitalium* was detected in 44/584 (8%, 95% CI 6, 10%) and vaginal dysbiosis in 375/584 (64%) of women. Median gestational age at delivery was 39 weeks +0 days (IQR 37+4, 40+1) in women with and 39 weeks +0 days (37+4, 40+0) in those without *M. genitalium*. In multivariable models, associations were not observed for any adverse birth outcomes. There was no interaction between *M. genitalium* and vaginal dysbiosis.

**Discussion:** *M. genitalium* in pregnancy was not associated with earlier gestational age at delivery or with other adverse birth outcomes. These findings do not support routine testing and treatment for *M. genitalium* in pregnancy.

## Introduction

*Mycoplasma genitalium* is a bacterium in the class Mollicutes, which is detected in 1 to 20% of pregnant women across different geographical settings^1–3^ and is more common among women with HIV infection than those without.^3^ *M. genitalium* infects endocervical epithelium as do *Chlamydia trachomatis* and *Neisseria gonorrhoeae*, which are reported to be associated with preterm birth, pregnancy loss, and other adverse pregnancy outcomes.^4,5^ Microorganism-induced inflammation is the hypothesised mechanism leading to the onset of preterm labour.^6,7^ Disturbance of the vaginal microbiota, including vaginal dysbiosis and bacterial vaginosis (BV), are also associated with both adverse pregnancy outcomes^8,9^ and with *M. genitalium*.^10^

The certainty of the body of existing evidence about the association between *M. genitalium* and adverse pregnancy outcomes remains limited by risks of bias in study design, analysis and reporting.^11^ Two systematic reviews, published in 2015^12^ and 2022,^11^ found the summary odds of preterm birth (<37 weeks gestation) were 1.9 times higher among women with *M. genitalium* than in those without. Meta-analysis was based on univariable data from six studies; two studies that included a multivariable analysis did not control for co-occurring STIs, *Candida* spp., or dysbiosis of the vaginal microbiota.

Being born prematurely is the single most important contributor to neonatal mortality, with no change in incidence in more than a decade in any world region.^13^ It is important to determine the role of infections such as *M. genitalium* to determine whether detection and treatment could improve outcomes. The South Africa is an upper middle-income country with large socio-economic disparities and high levels of HIV, STIs including *M. genitalium,*^3^ and BV^14^ in pregnancy, and preterm birth.^13^ The objectives of this study were to investigate the association between *M. genitalium* infection during pregnancy and gestational age at birth, preterm birth, miscarriage or stillbirth, and low birth weight, and the interaction between *M. genitalium* and vaginal dysbiosis.

## Methods

We followed a published protocol for this study^15^ and report it according to the Strengthening the Reporting of Observational Studies in Epidemiology guideline for cohort studies (supplementary material, file 1). The study received approval from the University of Cape Town Human Research Ethics Committee (reference: 676/2019) and from the local Department of Health (reference: EC_202010_017). The Canton of Bern Ethics Committee authorised analysis of de-identified data at the University of Bern (reference 2021-01209).

### Study design

We designed a prospective closed cohort study, which followed women enrolled during pregnancy until the first scheduled postnatal visit, 3-6 days after birth.^15^ The cohort study is part of a larger project called Philani Ndiphile (meaning ‘be healthy and I will be healthy’ in isiXhosa), which also includes a randomised controlled trial of screening and treatment strategies for STIs in pregnancy.^16^

### Study setting and population

The study was conducted at one public community health care centre in Buffalo City Metropolitan Municipality, Eastern Cape Province, South Africa. Enrolment took place from April 1, 2021 to August 29, 2023, with the last birth recorded on April 21, 2024; birth outcome data were sought until July 31, 2024. Trained research staff assessed women aged 18 years or older attending the antenatal clinic for eligibility and obtained their written consent for participation. After asking participants for the date of their last menstrual period, a trained study nurse ascertained gestational age by obstetric ultrasound. Initially, women whose pregnancy was <20 completed weeks were eligible. From September 2021, eligibility was extended to <27 weeks to increase enrolment during the COVID-19 pandemic and to align with another published study.^17^

### Study procedures and visits

At the enrolment visit, study staff entered social, demographic, clinical and behavioural information in a secure online database (Research Electronic Data Capture, REDCap, Vanderbilt University, TN). We assumed that all participants had been assigned female at birth and identified as women. A study nurse collected five vaginal swabs and two vaginal loop specimens. One swab was tested on-site with the Xpert CT/NG assay (GeneXpert, Cepheid, Sunnyvale, California, USA). Two swabs were placed in a single tube with digene Specimen Transport Medium (Qiagen, Hilden, Germany) and two dry flocked swabs were each placed in a separate tube (Copan, Brescia, Italy). Vaginal fluid specimens were smeared onto glass slides and air dried. Swab specimens were refrigerated at 4 degrees Celsius and glass slides were stored at room temperature at the clinic until weekly transport to the Department of Medical Microbiology, University of Pretoria within a maximum of 7 days. Women with a positive Xpert CT/NG result were offered same-day treatment for *C. trachomatis* and/or *N. gonorrhoeae*. Women with vaginal discharge but with negative results for *C. trachomatis* and *N. gonorrhoeae* received metronidazole and/or clotrimazole, according to South African guidelines.^18^

At a follow-up visit in the third trimester, scheduled at 30–34 weeks, we collected clinical and obstetric information and the same vaginal specimens, with repeat testing for *C. trachomatis* and *N. gonorrhoeae* and treatment if needed. A postnatal visit was scheduled three to six days after birth, following South African national guidelines. A study nurse recorded information reported by the woman and from the patient-held medical record. If the woman did not attend the visit, study staff collected the information from birth records and verified it through telephone interviews.

### Microbiological analyses

All laboratory analyses were done at the Department of Medical Microbiology, University of Pretoria. One dry flocked swab was stored at 4 degrees Celsius until extraction of genomic DNA (High Pure PCR Template Preparation kit, Roche Diagnostics, Mannheim, Germany), followed by storage at -20 degrees until after the end of the study period. We conducted PCR analyses on the LightCycler 480 II, Roche Diagnostics (Rotkreuz, Switzerland). using published primer and hydrolysis probe sequences and cycling conditions for detection and quantification of *M. genitalium, Trichomonas vaginalis*.^15^ For *Candida* spp. we used primer sequences from a laboratory-developed assay (Dr. Tania Crucitti, Institut Pasteur Madagascar). Vaginal smears were heat-fixed and Gram stained. Two trained people recorded Nugent scores^19^ independently. A third person examined slides with discrepant scores and a consensus was reached by discussion. We defined vaginal dysbiosis as a Nugent score of 4-10 and BV as Nugent score 7-10. All other swabs were stored for future analyses.

### Outcomes and statistical analysis

The primary outcome was gestational age at birth, measured in days. The continuous variable allowed description of the relationship between detection of *M. genitalium* and gestational age at delivery across the overall distribution and optimised statistical power. Secondary outcomes were preterm birth (<37 completed weeks of pregnancy), spontaneous abortion (<28 weeks) and stillbirth (≥28 weeks) and low birth weight (<2500 grams).^20^ We determined the sample size of the study for exposures with an assumed prevalence of 10% or more. Using Student’s t-test, 500–600 evaluable participants would provide 80% power to detect a difference of seven days (standard deviation, SD two days) in mean gestational age between groups with and without detection of the organism. Applying a Bonferroni correction, an alpha of 0·83% allowed for testing of six hypotheses.^15^

Statistical analyses followed a pre-specified plan (supplementary material, file 2). We used the median and interquartile range (IQR) to describe continuous variables and percentages and 95% confidence intervals (CI) for categorical variables in women with and without *M. genitalium* detected at enrolment. For each outcome, we fitted multivariable regression models. We controlled for pre-specified potential confounding variables measured at the baseline visit, based on published literature: age, education, alcohol consumption at baseline, previous preterm birth, living with HIV, detection of *C. trachomatis, N. gonorrhoeae*, *T. vaginalis* or *Candida* spp., and receipt of azithromycin by women who were treated for chlamydia. For the primary analysis, we included an interaction term for vaginal dysbiosis (Nugent 0-3 vs. Nugent 4-10). For the outcome of gestational age at birth, we used quantile regression to estimate marginal medians and their differences (with 95% CI),^21^ including all women with known birth outcome in the denominator. Quantile regression can account for skewed outcome distributions and assesses relationships across the outcome distribution, rather than focusing on the central tendency. For secondary outcomes, we used logistic regression to estimate marginal odds ratios and risk differences. The model for preterm birth only included women with a live birth in the denominator to avoid the competing risk of pregnancy loss before 37 weeks. We combined pregnancy loss from spontaneous abortion and stillbirth because of the small number of stillbirths. For all regression-based analysis, we excluded women with a missing birth outcome. We used multiple imputation with chained equations to impute missing values in pre-specified potential confounding variables (<2% for relevant variables). We confirmed the results in an analysis using cases with complete data (supplementary material 3).

We conducted four post-hoc sensitivity analyses. We repeated analyses: using *M. genitalium* test results from the third trimester visit as the exposure; excluding women who had co-infection with *C. trachomatis, N. gonorrhoeae* or *T. vaginalis*; excluding women who received treatment for chlamydia with azithromycin; and with birth weight as a continuous outcome. Statistical analyses were done in Stata (version 18.0) and R (version 4.4.2).

## Results

We enrolled 603 pregnant women (of 846 assessed, 71%, Figure 1). Of 586 eligible included women, 41 experienced the end of their pregnancy before the scheduled third trimester visit and, of the remaining 545 women, 440 (81%) attended both visits. We analysed data from 584/586 women (one woman had a missing *M. genitalium* test result at enrolment and one withdrew before her birth outcome was known). Information about the primary or secondary birth outcomes was known for 560/603 enrolled (93%).

**Figure 1:**
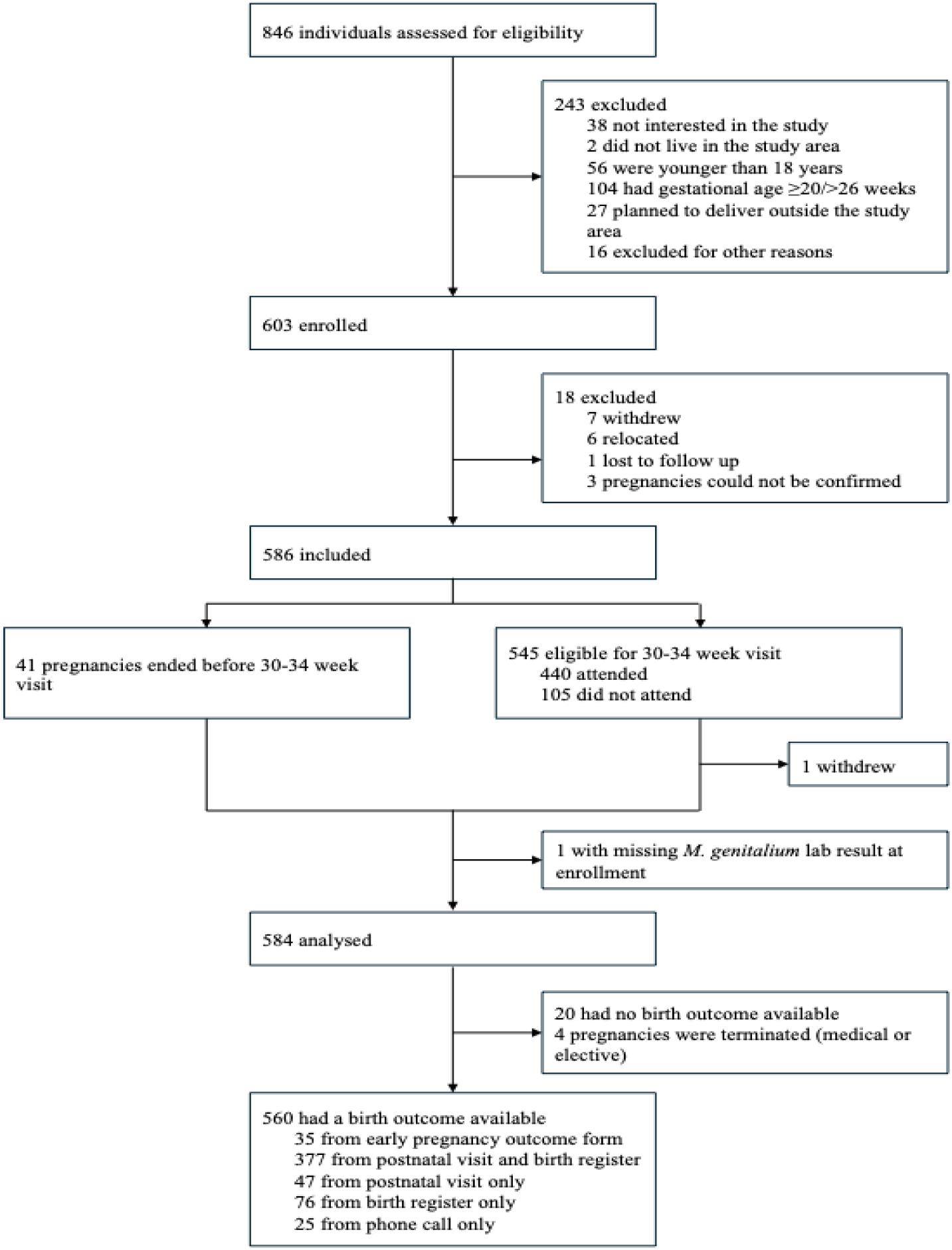
Participant flow in the Philani Ndiphile cohort study.

The median age at enrolment was 28 years (IQR 24, 33) and the median gestational age was 13 weeks +5 days (10+0, 19+1) (Table 1). At enrolment, 159/584 (27%) women were living with HIV, with most on antiretroviral therapy (ART), syphilis was detected in 16/584 (3%), and 28 (5%) reported at least one prior preterm birth. *C. trachomatis* was detected in 91 (16%) of women, *N. gonorrhoeae* in 33 (6%), *T. vaginalis* in 61 (10%) and *Candida* spp. in 203 (35%). There were 375 (64%) women with vaginal dysbiosis (Nugent score 4-10) and 314 (54%) with BV (Nugent score 7-10).

**Table 1:** Characteristics of women, overall and according to results of testing for *M. genitalium* at enrolment.

| Characteristic | Total (n=584) | *M. genitalium* test negative (n=540) | *M. genitalium* test positive (n=44) | p value |
| --- | --- | --- | --- | --- |
| Gestational age at enrolment, weeks+days | 13+5 (10+0, 19+1) | 13+5 (10+0, 19+1) | 14+5 (10+4, 19+1.5) | 0.63 |
| Missing, n | 2 | 1 | 1 |  |
| Age | 28 (24, 33) | 28 (24, 33) | 27 (23, 31) | 0.29 |
| Educational level |  |  |  | 0.56 |
| Less than grade 10 | 37 (6%) | 33 (6%) | 4 (9%) |  |
| Grade 10 or 11 | 187 (32%) | 175 (32%) | 12 (27%) |  |
| Grade 12 or higher | 360 (62%) | 332 (61%) | 28 (64%) |  |
| Any alcohol consumption |  |  |  | 0.33 |
| No | 428 (73%) | 399 (74%) | 29 (66%) |  |
| Yes | 156 (27%) | 141 (26%) | 15 (34%) |  |
| Living with HIV |  |  |  | 0.02 |
| No | 425 (73%) | 401 (74%) | 24 (55%) |  |
| Yes, on ART | 121 (21%) | 106 (20%) | 15 (34%) |  |
| Yes, not on ART | 38 (7%) | 33 (6%) | 5 (11%) |  |
| Syphilis |  |  |  |  |
| No | 568 (97%) | 526 (97%) | 42 (95%) | 0.34 |
| Yes | 16 (3%) | 14 (3%) | 2 (5%) |  |
| At least one prior preterm birth |  |  |  | 1.00 |
| No | 545 (95%) | 503 (95%) | 42 (95%) |  |
| Yes | 28 (5%) | 26 (5%) | 2 (5%) |  |
| Missing | 11 | 11 | 0 |  |
| Vaginal dysbiosis |  |  |  | 0.16 |
| Negative (Nugent 0-3) | 209 (36%) | 198 (37%) | 11 (25%) |  |
| Positive (Nugent 4-10) | 375 (64%) | 342 (63%) | 33 (75%) |  |
| Nugent score | 7 (1, 9) | 7 (0, 9) | 8 (4.5, 10) | 0.03 |
| *C. trachomatis* |  |  |  | <0.01 |
| Negative | 493 (84%) | 464 (86%) | 29 (66%) |  |
| Positive | 91 (16%) | 76 (14%) | 15 (34%) |  |
| *N. gonorrhoeae* |  |  |  | 0.17 |
| Negative | 551 (94%) | 512 (95%) | 39 (89%) |  |
| Positive | 33 (6%) | 28 (5%) | 5 (11%) |  |
| *T. vaginalis* |  |  |  | <0.01 |
| Negative | 523 (90%) | 492 (91%) | 31 (70%) |  |
| Positive | 61 (10%) | 48 (9%) | 13 (30%) |  |
| *Candida* spp. |  |  |  | 0.5 |
| Negative | 376 (65%) | 350 (65%) | 26 (59%) |  |
| Positive | 203 (35%) | 185 (35%) | 18 (41%) |  |
| Missing | 5 | 5 | 0 |  |
| Azithromycin treatment for *C. trachomatis* during pregnancy |  |  |  | 0.02 |
| No | 457 (78%) | 429 (79%) | 28 (64%) |  |
| Yes | 127 (22%) | 111 (21%) | 16 (36%) |  |

Data are n (%) or median (interquartile range, IQR)
ART, antiretroviral therapy; HIV, human immunodeficiency virus

At enrolment, 44/584 women (8%, 95% CI 6, 10%) tested positive for *M. genitalium* (Table 1). Proportions of women with a positive test result for *M. genitalium* were higher among women living with HIV, women with positive test results for *C. trachomatis*, *T. vaginalis*, and women who had received azithromycin following a positive test result for *C. trachomatis*. The median Nugent score was higher 8 (4·5, 10) in women with *M. genitalium* and 7 (0, 9) in those without.

The distribution of gestational age at delivery was right-skewed with median 39 weeks (37+4, 40+1) (Table 2, Figure S1). Gestational age at birth was earlier for women aged 35 years and older than for younger women (Table S1). Of 564 women with a known birth outcome, 114 (20%) gave birth preterm, and 72 (14%) of newborns were of low birth weight. Gestational age outcomes were missing for 24 women (4%), including four women with an elective or medically indicated termination, with similar patterns of missingness for preterm birth, live birth, and pregnancy loss.

**Table 2:**
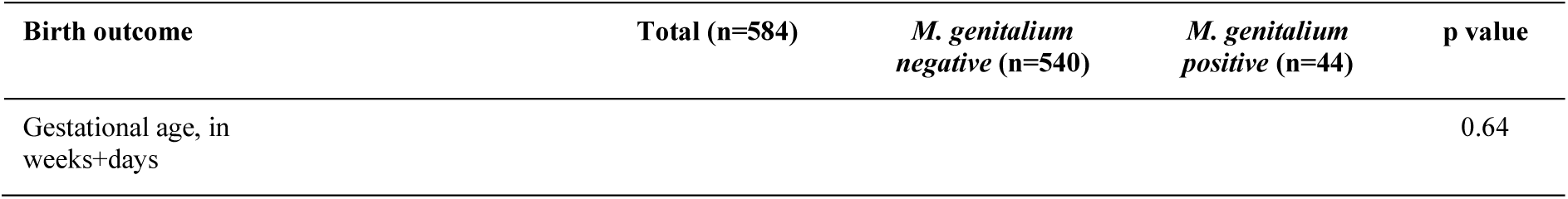
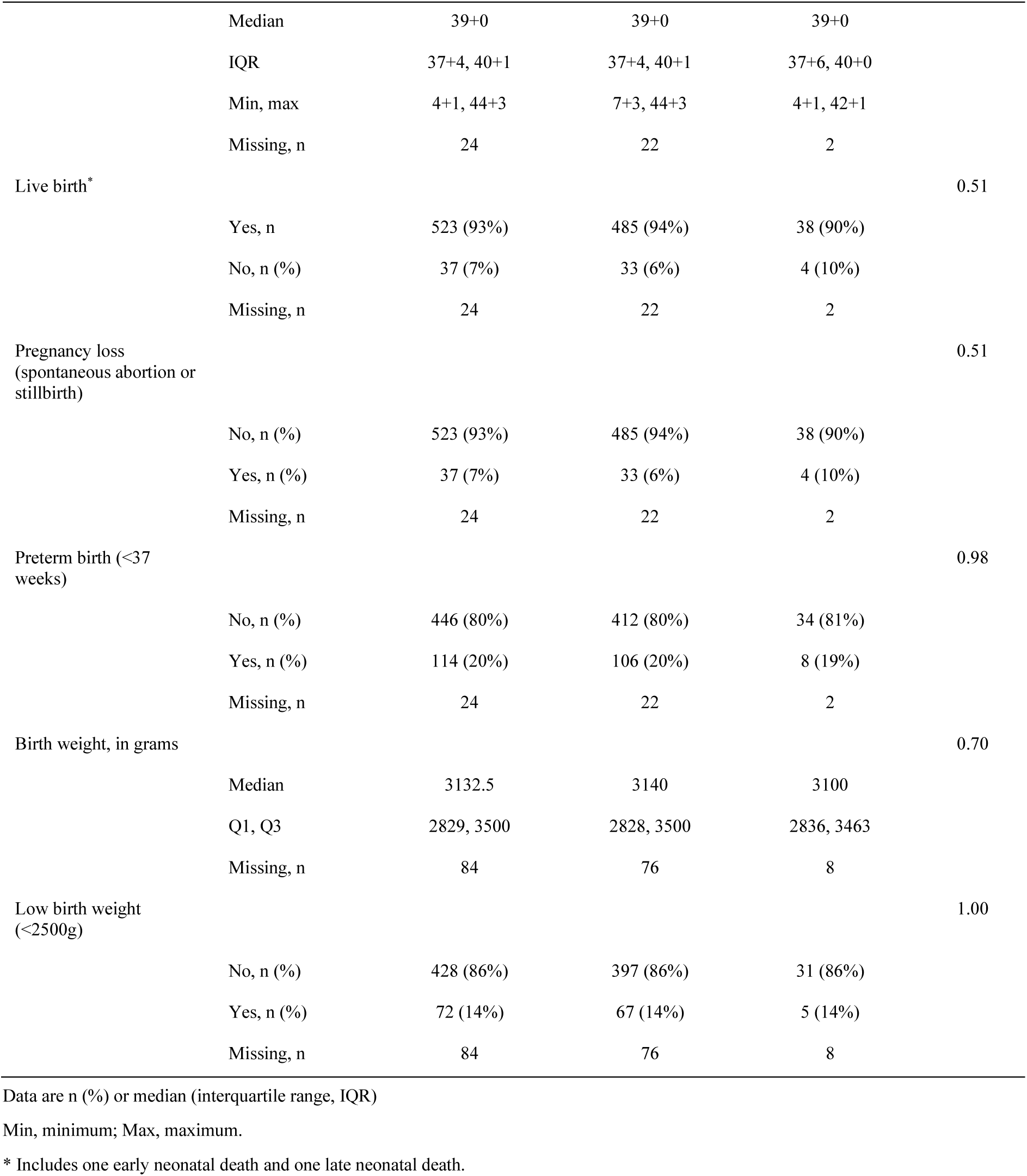
*M. genitalium* and associations with gestational age at birth, preterm birth and other secondary outcomes.

| Birth outcome | Total (n=584) | *M. genitalium*<br>negative (n=540) | *M. genitalium*<br>positive (n=44) | p value |
| --- | --- | --- | --- | --- |
| Gestational age, in weeks+days |  |  |  | 0.64 |

|  |  |  |  |  |  |
| --- | --- | --- | --- | --- | --- |
|  | Median | 39+0 | 39+0 | 39+0 |  |
|  | IQR | 37+4, 40+1 | 37+4, 40+1 | 37+6, 40+0 |  |
|  | Min, max | 4+1, 44+3 | 7+3, 44+3 | 4+1, 42+1 |  |
|  | Missing, n | 24 | 22 | 2 |  |
| Live birth* |  |  |  |  | 0.51 |
|  | Yes, n | 523 (93%) | 485 (94%) | 38 (90%) |  |
|  | No, n (%) | 37 (7%) | 33 (6%) | 4 (10%) |  |
|  | Missing, n | 24 | 22 | 2 |  |
| Pregnancy loss<br>(spontaneous abortion or<br>stillbirth) |  |  |  |  | 0.51 |
|  | No, n (%) | 523 (93%) | 485 (94%) | 38 (90%) |  |
|  | Yes, n (%) | 37 (7%) | 33 (6%) | 4 (10%) |  |
|  | Missing, n | 24 | 22 | 2 |  |
| Preterm birth (<37<br>weeks) |  |  |  |  | 0.98 |
|  | No, n (%) | 446 (80%) | 412 (80%) | 34 (81%) |  |
|  | Yes, n (%) | 114 (20%) | 106 (20%) | 8 (19%) |  |
|  | Missing, n | 24 | 22 | 2 |  |
| Birth weight, in grams |  |  |  |  | 0.70 |
|  | Median | 3132.5 | 3140 | 3100 |  |
|  | Q1, Q3 | 2829, 3500 | 2828, 3500 | 2836, 3463 |  |
|  | Missing, n | 84 | 76 | 8 |  |
| Low birth weight<br>(<2500g) |  |  |  |  | 1.00 |
|  | No, n (%) | 428 (86%) | 397 (86%) | 31 (86%) |  |
|  | Yes, n (%) | 72 (14%) | 67 (14%) | 5 (14%) |  |
|  | Missing, n | 84 | 76 | 8 |  |

Data are n (%) or median (interquartile range, IQR)
Min, minimum; Max, maximum.
\* Includes one early neonatal death and one late neonatal death.

In unadjusted analyses, the median gestational age was 39+0 weeks for women with and without positive test results for *M. genitalium* and the frequencies of all other measured birth outcomes were similar between the two groups (Table 2, Figure S2). The distribution of gestational age at birth was similar for women with and without any of the other organisms assessed or vaginal dysbiosis at enrolment (Table S1, Figure S2). Results were similar in post-hoc sensitivity analyses, when restricted to women with *M. genitalium* but without any other STI (Figure S3) and to women who did not receive treatment with azithromycin (Figure S4).

In multivariable analysis among women with vaginal dysbiosis, gestational age at birth was slightly earlier among women with *M. genitalium* than without (marginal median difference - 4 days, 95% CI -1+2, 0+1, Table S3). Among women without vaginal dysbiosis, gestational age at birth was 6 days longer (-0+2, 2+0 weeks) in women with *M. genitalium* than without (Figure 2, Table S3). The confidence intervals for the difference in both groups of women included zero. Findings were unchanged for analyses restricted to women with live births (Table S4).

**Figure 2:**
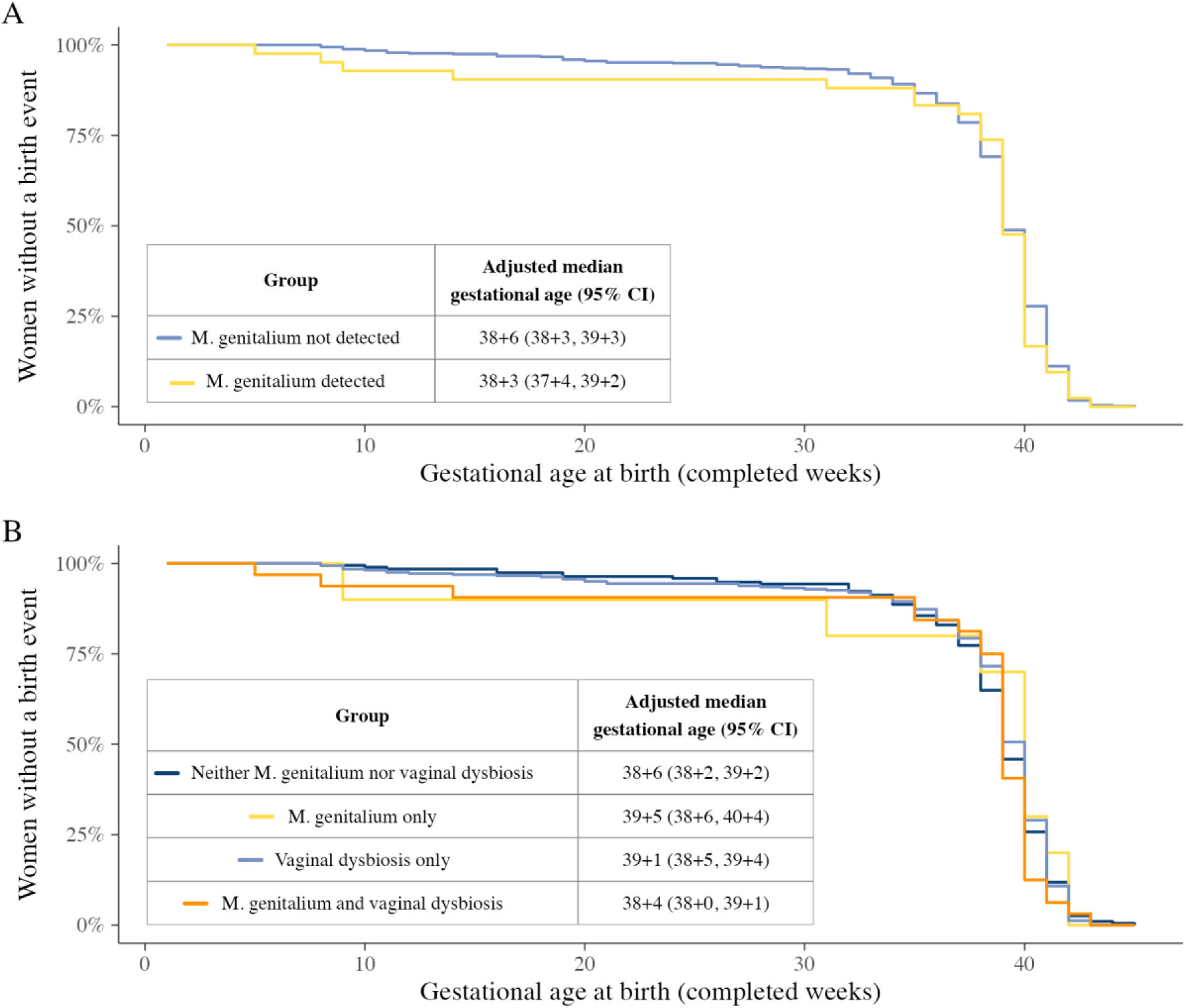
Time to birth event by *M. genitalium* detection and vaginal dysbiosis status. Survival curves showing the proportion of women without a birth event across gestational age. (A) Stratified by *M. genitalium* detection at baseline. (B) Stratified by *M. genitalium* and vaginal dysbiosis detection at baseline. Tables show adjusted marginal median gestational ages with 95% confidence intervals, estimated with median regression and adjusted for: age, education, alcohol consumption at baseline, previous preterm births, living with HIV, receipt of azithromycin, *C. trachomatis*, *T. vaginalis*, *N. gonorrhoeae*, *Candida* spp. and vaginal dysbiosis. The regression model for panel B includes an interaction term between *M. genitalium* and vaginal dysbiosis, the model for panel A does not.

We report the results for secondary birth outcomes without stratification by vaginal dysbiosis because there was no apparent interaction. Outcomes were similar for women with and without *M. genitalium* detected (Table 3). Among all participants with a known birth outcome, adjusted estimates of median gestational age were 38+3 weeks (95% CI 37+5, 39+1) in women with *M. genitalium* and 38+6 (38+2, 39+2) in women without. Marginal odds ratios for *M. genitalium* test positivity were 1·74 for pregnancy loss (0·56, 5·42), 1·04 for preterm birth (0·45, 2·4), and 0·95 for low birth weight (0·34, 2·62). Differences in all measured birth outcomes in women with and without *M. genitalium* were close to zero (Table 3).

**Table 3:** Adjusted estimates of associations between *M. genitalium* and birth outcomes.

| Outcome |  | Comparison* | Estimate |
| --- | --- | --- | --- |
| Gestational age, in completed weeks+days | All women | *M. genitalium* not detected | 38+6 (38+2, 39+2) |
|  |  | *M. genitalium* detected | 38+3 (37+5, 39+1) |
|  |  | Median difference | -0+3 (-1+0, 0+2) |
|  | Women with live births | *M. genitalium* not detected | 38+5 (38+2, 39+2) |
|  |  | *M. genitalium* detected | 38+5 (38+1, 39+3) |
|  |  | Median difference | 0+0 (-0+4, 0+4) |
| Pregnancy loss | All women | Marginal odds ratio | 1.74 (0.56, 5.42) |
|  |  | Risk difference | 0.02 (-0.03, 0.07) |
| Preterm birth | All women | Marginal odds ratio | 1.04 (0.45, 2.4) |
|  |  | Risk difference | 0.01 (-0.11, 0.13) |
|  | Women with live births | Marginal odds ratio | 0.75 (0.25, 2.28) |
|  |  | Risk difference | -0.03 (-0.15, 0.09) |
| Birth weight, in grams | All women | *M. genitalium* not detected | 3060.64 (2878.67, 3242.62) |
|  |  | *M. genitalium* detected | 3023.54 (2695.62, 3351.45) |
|  |  | Median difference | -37.11 (-335.89, 261.67) |
|  | Women with live births | *M. genitalium* not detected | 3060.05 (2880.77, 3239.33) |
|  |  | *M. genitalium* detected | 3037.83 (2707.47, 3368.19) |
|  |  | Median difference | -22.22 (-323.48, 279.03) |
| Low birth weight | All women | Marginal odds ratio | 0.95 (0.34, 2.62) |
|  |  | Risk difference | -0.01 (-0.13, 0.12) |
|  | Women with live births | Marginal odds ratio | 0.98 (0.35, 2.71) |
|  |  | Risk difference | 0 (-0.14, 0.13) |

\* For gestational age, contrasts are marginal medians and median differences. For other birth outcomes, contrasts are marginal odds ratios and risk differences. Estimates are adjusted for: age, education, alcohol consumption at baseline, previous preterm births, living with HIV, receipt of azithromycin, and exposure to vaginal dysbiosis, *C. trachomatis*, *T. vaginalis*, *N. gonorrhoeae* and *Candida* spp.

In post-hoc analysis, results were similar when we repeated the analysis with exposure at the third trimester visit (Table S5). Birth weight was similar in women with and without *M. genitalium* (median difference -37.11 g, 95% CI -335.89, 261.67) (Table 3, Figure S5).

## Discussion

In a South African antenatal cohort with ultrasound-confirmed dating and high ascertainment of pregnancy outcomes, the prevalence of *M. genitalium* was 8% (95% CI 6-10). Detection of *M genitalium* at enrolment was not associated with earlier gestational age at birth. This result did not change in multivariable analyses, controlling for multiple co-occurring organisms, and there was no interaction between *M. genitalium* and vaginal dysbiosis. Secondary outcomes showed effect estimates close to the null, although confidence intervals were wide for rarer outcomes.

### Strengths and weaknesses of study design

The main strengths of this study were its prospective design, with prespecified objectives and analysis plan,^15^ accurate measurement of gestational age at enrolment and high ascertainment of birth outcomes. By choosing gestational age at birth as our primary outcome, we were able to examine the relationship between *M. genitalium* infection and the full distribution of gestational age. Other strengths include testing for other STIs and *Candida* spp. Our study also has limitations. First, women who tested positive for *C. trachomatis* received azithromycin, which would cure *M. genitalium* amongst those with co-infection because the prevalence of mutations associated with macrolide resistance is low in South Africa.^22^ Our findings did not change, however, when restricting to women who did not receive azithromycin. Second, the cohort enrolled women when they presented for antenatal care so very early pregnancies and pregnancy losses are underrepresented. A median of 13 weeks of gestation at enrolment is early for a study in a resource-constrained setting, however. Third, there are differences over the course of pregnancy in the non-infectious causes of pregnancy loss and preterm birth; we assumed that these would not differ between women with and without *M. genitalium* and we did not control for them. Fourth, enrolment at a single clinic may limit the generalisability of our findings.

### Comparison with other studies

In our study among pregnant women in South Africa, the median gestational age at birth was the same in women with and without *M. genitalium* in both univariable and multivariable analyses. We found two studies from the USA that reported on gestational age at birth as a secondary outcome.^2,23^ One study (N=281), reported mean gestational age at birth 38.5 weeks in women with *M. genitalium* (with or without another STI) and 38.5 weeks in women with no STI.^2^ In multivariable analysis, the adjusted mean difference was 0.16 weeks (95% CI -0.17, 1.03). In the other study (N=81), median gestational age at birth was 39+0 weeks in women with *M. genitalium* and those without; there were, however, only 7 women with *M. genitalium*.^23^ For the dichotomised outcome of preterm birth, we found two other studies published after the end of the search dates of the systematic reviews. In South Africa, authors reported an adjusted odds ratio of 2.1 (0.6 to 7.5) in a secondary analysis of a study among women without HIV infection.^24^ In India, authors found a univariable odds ratio of 3.4 (1.1 to 10.4).^25^

Studies reporting on *M. genitalium* and spontaneous abortion or miscarriage have used gestational age cut-offs from 16 to 24 weeks, or did not report a cut-off. In a meta-analysis of 6 studies the odds ratio was 1.0 (0.5 to 1.9).^11^ In a study in Denmark, published in 2026, authors found 0.9% of women with *M. genitalium* and no woman with spontaneous pregnancy loss before 22 weeks.^26^ For outcomes related to birth weight, we found similar values in women with and without *M. genitalium* in both univariable and multivariable analyses. Our findings were consistent with the only other study reporting on both outcomes^23^ at the time of the most recent systematic review.^11^ In four studies published after the end of the search dates of the systematic reviews, findings were mixed. Three studies found higher odds of low birth weight^24^ or lower mean birthweight^2,10^ in multivariable analysis. In the study in Papua New Guinea, there was little difference in crude birth weight for women with and without *M. genitalium* (0.7g, 95% CI -135 to 137g). In multivariable analyses, a difference was found -167g (95% CI -324 to -10g), but reasons for negative confounding were not explored.^10^ In India, low birth weight was not associated with *M. genitalium* detection in univariable analysis.^25^

### Meaning of the study

Our study findings highlight incomplete understanding of the relationship between *M. genitalium* throughout pregnancy and adverse pregnancy outcomes.^27^ Whilst there is evidence that infection-induced inflammatory cytokines trigger a pathological cascade of signalling leading to early onset of labour,^6,28^ the inflammatory potential of *M. genitalium* is not clear. In cultured vaginal and cervical epithelial cells infected with *M. genitalium*, secretion of proinflammatory cytokines can be detected.^29^ In a clinical study among non-pregnant young women, levels of inflammatory cytokines and chemokines were not, however, elevated among those with *M. genitalium*.^30^ We could not directly compare outcomes between STIs in our study because women with *C. trachomatis* or *N. gonorrhoeae* received treatment and we did not measure inflammatory markers. It is possible that the small effect sizes in our study for any outcome reflect, in part, the prespecified objectives and efforts to reduce confounding and risks of selection, measurement and outcome reporting biases. We did not find prespecified analysis plans for the other studies.

Vaginal dysbiosis was common in our study population, in common with studies among women in southern Africa.^14^ We did not find convincing evidence that vaginal dysbiosis modified the effect of *M. genitalium* on gestational age at birth, despite earlier studies suggesting this possibility.^31^ The finding that, among women without vaginal dysbiosis, those with *M. genitalium* had a slightly longer gestational age at birth than those without is compatible with chance. A study from Denmark found no association between vaginal microbial communities, pregnancy loss and genital mycoplasmas, including *M. genitalium*.^26^ We found the distributions of gestational age at delivery to be similar for all measured organisms (Figure S2).

### Implications for further research and clinical practice

Our study highlights a number of important research needs. First, additional research with prespecified analysis plans about *M. genitalium* and its association with gestational age at delivery and other adverse pregnancy outcomes is needed to improve reproducibility. Second, such studies should use a holistic approach to measuring the presence of different STIs and vaginal microbiota with state-of-the-art diagnostic techniques, including rRNA sequencing.^15^ Third, measurement of inflammatory markers and quantitation of organism load are needed to determine whether the overall levels of inflammation are more important than individual species in the prediction of preterm birth. A randomised controlled trial of the effect of antenatal screening for *C. trachomatis*, *N. gonorrhoeae*, *T. vaginalis* and BV in Papua New Guinea^17^ found no reduction in a composite outcome of preterm birth or low birthweight or both. Stored samples from these trials could be tested for *M. genitalium* to determine the causal association. The association between *M. genitalium* and adverse birth outcomes requires further research to clarify the potential benefits of screening and treatment interventions for *M. genitalium* during pregnancy. At present, there is insufficient evidence to support routine screening for *M. genitalium* in pregnancy to prevent preterm birth.

## Data Availability

All data produced in the present study are available upon reasonable request to the authors

## Contributors

Conceptualisation: RMSG, JHHMvdW, RPHP, NL; Data curation and full access to the dataset: RMSG, LB-M, J-BR; Formal analysis: RMSG, LB-M, J-BR; Funding acquisition: RMSG, AM-M, JDK, JHHMvdW, RPHP, NL; Investigation: RMSG, MMM, HJ, EM, NL; Methodology: RMSG, HJ, LB-M, J-BR, JHHMvdW, RPHP, NL; Project administration: RMSG, MMM, RPHP, NL; Resources: RMSG, MMM, HJ, LB-M, J-BR; Software and visualisation: LB-M, J-BR; Supervision: JHHMvdW, RPHP, NL; Validation: RMSG, MMM, HJ, LB-M, J-BR; Writing – original draft: RMSG, LB-M, NL; Writing – reviewing and editing: all authors.

## Declaration of interests

The Foundation for Professional Development received GeneXpert machines through a contract between the University of Southern California and Cepheid. All authors declare no conflicts of interest.

## Data sharing

Deidentified individual participant data that underly the results reported in this article will be made available to researchers who provide a methodologically sound proposal. Proposals should be directed to the corresponding author, and data requesters will need to sign a data access agreement.

## Acknowledgements

The authors thank the participants of the Philani Ndiphile study.

## Financial support

This study was supported by the United States National Institute of Allergy and Infectious Diseases (grant number R01AI149339 to AM-M, JDK) and the Swiss National Science Foundation (project numbers 191225 to RMSG and 197831 to JHHMvdW, RPHP, NL). RMSG, MMM, HJ, CMB were employed for the duration of this study.

## Ethics approval

This study received approval from the University of Cape Town’s Human Research Ethics Committee (Reference: 676/2019) and from the local Department of Health (Reference: EC_202010_017). Authorisation to analyse de-identified data at the University of Bern has been granted by the Canton of Bern Ethics Committee (Reference 2021-01209).

## Notes

### Clinical Protocols

https://www.ncbi.nlm.nih.gov/pubmed/38154893

### Author Declarations

The Human Research Ethics Committee of the University of Cape Town (reference: 676/2019) and the Department of Health East London (reference: EC_202010_017) gave ethical approval for this work. The Canton of Bern Ethics Committee authorised analysis of de-identified data at the University of Bern (reference 2021-01209).

