## supplementary material, file 2 for "*Mycoplasma genitalium* infection and adverse pregnancy outcomes among pregnant women in South Africa: prospective cohort study"

### Statistical Analysis Plan (SAP)

#### Administrative Information

| | |
|---|---|
| Project number: | 1855 |
| Trial registration number: | - |
| SAP version: | 3.0 (21.02.2025) |
| Protocol version: | 1.0 (06.12.2023) |

#### Contributors

| Name | Affiliation | Role in SAP writing |
|---|---|---|
| Dr. Jean-Benoît Rossel | CTU Bern | Author |
| Prof. Nicola Low | ISPM Bern | Sponsor-Investigator |
| Dr. Lydia Braunack-Mayer | ISPM Bern | Researcher |
| Dr. Ranjana Gigi | ISPM Bern and Foundation for Professional Development | Researcher |
| Dr. Lukas Bütikofer | CTU Bern | Reviewer |

#### Approved by

| Name | Affiliation | Study Role | Date and Signature (wet ink) |
|---|---|---|---|
| Dr. Jean-Benoît Rossel | CTU Bern | Trial Statistician | |
| Dr. Lukas Bütikofer | CTU Bern | Senior Statistician | |
| Prof. Nicola Low | ISPM Bern | Sponsor-Investigator | |

#### Revision history

| Version | Chapter / Section | Summary of changes |
|---|---|---|
| 1.0 | | First version |
| 2.0 | 2.2.3 | • Correction of three-category education, and addition of a binary version (11 and less vs. 12 and above).<br>• Addition of a new variable `prev_mis_stillb_control` for "Any previous miscarriages or stillbirths" |
| | 3.1 | New definition of the FAS: exclude patients with missing information about baseline exposure. |
| | 3.2.2 | Effects will be presented separately; one effect for women with bacterial vaginosis, and one effect for women without it. |
| | 3.2.3 | • Consideration of a residuals-versus-fits plot.<br>• Analysis of gestational age with ordinal logistic regression: add the possibility to use tertiles (i.e., three categories of gestational age) and multinomial logistic regression if the proportional odds assumption is violated. |
| | 3.2.4 | • Miscarriages and stillbirths will always be considered together in the analysis. The multinomial model (live birth vs. miscarriage vs. stillbirth) is then replaced by a classical logistic regression model (live birth vs. miscarriage/stillbirth).<br>• We also add the possibility to reduce the number of covariates, in case of numerical issues. |
| | 3.2.7 | New paragraph about sensitivity analysis |
| 3.0 | Contributors | Dr. Lydia Braunack-Mayer added to the list of contributors. |
| | 2.2.3, 3.2.1, 3.2.2 | New definition of vaginal dysbiosis: Nugent score $\geq$ 4. |
| | 3.2.3 | Quantile regression and survivial analysis will be considered as alternatives to linear regression models. |
| | 3.3 | • Addition of R version 4.2.2 (or more recent) to the list of statistical software used to perform analyses.<br>• Acknowledgement that AI-based tools, such as GitHub Copilot, may be used to support code development. |
| | 3.4 | Corrections to terminology of Nugent score categories |

### 1. Introduction

#### 1.1 Background and rationale

The prevalence of sexually transmitted infections (STIs), such as those caused by *Chlamydia trachomatis, Neisseria gonorrhoeae* and *Trichomonas vaginalis,* is high among pregnant women in South Africa and most infections remain untreated [1-3]. Adverse outcomes of pregnancy, including preterm birth, are also common among women in South Africa. Numerous studies have shown that STIs, colonization with other vaginal microorganisms, or a combination of these during pregnancy have associations with adverse pregnancy and birth outcomes [4-7]. *Mycoplasma genitalium* is a sexually transmissible bacterium, for which less is known than for other STIs about its association with adverse pregnancy outcomes [8]. These associations are most often found in univariable analyses, with few studies investigating the role of confounding.

A cohort study was designed to investigate associations between the presence of lower genital tract organisms in pregnancy and adverse pregnancy outcomes [9].

#### 1.2 Objectives

The overall aim of the cohort study is to investigate associations between the presence of lower genital tract organisms in pregnancy and adverse pregnancy outcomes, as described in the cohort study protocol [7].

The specific objective of this analysis is to investigate the association between the presence of *M. genitalium* in pregnancy and adverse pregnancy outcomes.

### 2. Data management

#### 2.1 Data export

Data are exported from a REDCap database and were prepared in advance. A csv-file will be available for the analysis.

#### 2.2 Data preparation

A codebook is available with the original REDCap database. Here are other important variables in the csv-file.

##### 2.2.1 Outcomes

| Concept | Variable or Derivation | Type |
|---|---|---|
| Primary outcome: Gestational age at birth, in days. | `outcome_GA` | Numerical |
| Secondary outcome: preterm birth (<37 completed weeks of gestation) | `outcome_GA < 259` | Binary |
| Secondary outcome: low birth weight | `pn_birth_weight_b1 < 2500 OR pn_birth_weight_b2 < 2500` | Binary |
| Secondary outcome: Miscarriage | `outcome_b1 == 3 and outcome_GA < 196` | Binary |
| Secondary outcome: Stillbirth | `outcome_b1 == 3 and outcome_GA > 195 and outcome_GA!=.` | Binary |

##### 2.2.2 Exposure

*M. genitalium* is the primary exposure, detected by PCR. Vaginal samples were taken at two antenatal visits and stored, with testing by PCR after the end of the enrolment and follow-up period. *M. genitalium* will be coded as a binary variable (positive or negative) in two different ways: 1) at enrollment only, and 2) at any time among enrollment and a second visit in the third trimester of pregnancy.

##### 2.2.3 Potential confounding variables and other variables to examine in descriptive analysis

| Variable | Type | Concept |
|---|---|---|
| calculated_age | Numerical | Age, in years |
| sd_education | Categorical | Educational level, in 3 categories (0: less than grade 10, 1: grade 10 or 11, 2: grade 12 or above).<br>A binary version is also possible (0-1, 2). |
| bq_drugs_during_pregnancy___1 | Binary | Alcohol consumption at baseline |
| w32_drugs_used___1 | Binary | Alcohol consumption at 30-34 week follow-up visit |
| bq_hiv_status | Categorical | HIV infection at baseline, in three categories: 1: HIV negative, 2: HIV positive on ART, 3: HIV positive, not on ART. |
| w32_hiv_status | Categorical | HIV infection at 30-34 week follow-up visit |
| bq_delivery_timing1 | Numerical or binary | Number of prior preterm births (also possible to code as 0 (no) and >0 (yes)). |
| prev_mis_stillb_control | Binary | Any previous miscarriages or stillbirths |
| sti_result_tv | Binary | *T. vaginalis* samples taken and tested after giving birth. |
| sti_result_ct | Binary | *C. trachomatis* samples taken and tested at two antenatal visits. Treated if result positive. |
| sti_result_ng | Binary | *N. gonorrhoeae* samples taken and tested at two antenatal visits. Treated if result positive. |
| sti_result_nugent | Numerical | Nugent score (value between 0 and 10), from which we can define vaginal dysbiosis as a binary variable (yes if Nugent score ≥ 4). Alternative, three categories (0-3, 4-6, 7-10). |
| w32_nugent_res | Numerical | Nugent score at 30-34 week follow-up visit |
| sti_treatment___1 | Binary | Azithromycin exposure |

### 3. Analysis

#### 3.1 Patient sets

Two different patient sets will be considered in our analyses:

- The Full Analysis Set (FAS) contains all women from the cohort study, that are either positive or negative to the exposure at baseline. Women with missing information about *M. genitalium* at baseline are excluded.
- The Live Birth Set (LBS) is a subset of the FAS, including women who had a live birth (includes babies previously coded as early neonatal death). Women with termination and those with miscarriage/stillbirth (according to the `outcome_b1` variable) do not belong to this LBS.

#### 3.2 Analysis methods

##### 3.2.1 Descriptive analysis

We start by describing the exposure and the potential confounding variables for the women in the FAS. In general, categorical variables will be summarized with raw numbers and percentages, and continuous variables will be summarized with median, interquartile range and total range (and displayed in histograms). For variables measured at baseline and 30-34 weeks, we will summarize findings for each visit separately.

We first present these descriptive statistics for the entire FAS. In a second step, we compare the exposed and non-exposed women. Categorical variables are compared via chi-squared tests or Fisher's exact tests in case of small numbers (less than 5) in some cells of the contingency tables. Continuous variables are compared via Mann-Whitney-Wilcoxon tests.

We also present the outcomes in the same way, on the FAS. In case of a termination (according to the `outcome_b1` variable), all outcomes are considered as missing.

Finally, we further explore some data. Nugent score is measured at baseline and at 30-34 week follow-up visit. We will do some cross-tables between these two measurements, by considering the binary version (0-3, 4-10) or the three-category version (0-3, 4-6, 7-10). These measures, as well as the azithromycin exposure, will then be cross-tabled with *C. trachomatis* and *N. gonorrhoeae* results.

##### 3.2.2 General principles for the regression models

For each outcome, we will fit multivariable regression models with the outcome as dependent variable, and the baseline exposure and the following confounders as predictors (also measured at baseline): age, educational level, alcohol consumption, HIV infection, and prior preterm birth. We will also control for the presence of *C. trachomatis, N. gonorrhoeae, T. vaginalis* and vaginal dysbiosis (Nugent score 4–10). We may also add an interaction between the exposure and bacterial vaginosis. To add some flexibility to the model, continuous confounders will be added using the best fitting fractional polynomial of grade two (e.g., using `fp` in Stata).

For continuous outcomes, we will estimate mean differences (i.e., regression coefficients), while for categorical outcomes, we will report marginal odds ratios and risk differences. All effect measures will be reported with 95% confidence intervals and p-values based on robust standard errors (e.g. using option `vce(robust)` in Stata). Due to the presence of the interaction between the exposure and vaginal dysbiosis in the models, we will report one effect for women with vaginal dysbiosis and one effect for women without vaginal dysbiosis.

In case of a termination (according to the `outcome_b1` variable), all outcomes are considered as missing (see section 3.2.5). We will show results on both FAS and LBS.

##### 3.2.3 Analysis of the gestational age

Gestational age is a continuous outcome, and its distribution is peaked (leptokurtotic) and slightly left-skewed (Annex, figure 1). Classical linear regression based on normality assumption may fail to model it properly. We will try it anyway and visually assess the normality assumption of the residuals with a quantile-quantile plot and a residuals-versus-fits plot. If necessary, the following alternative approaches will be attempted: quantile regression; survival analysis, or; generalized linear models with other distributions (e.g., Gamma). Another option would be to define categories based on quintiles or tertiles, and then use ordinal logistic regression. If the proportional odds assumption is violated, multinomial logistic regression may be considered.

##### 3.2.4 Analysis of the birth outcome

We want to compare live birth to the two other outcomes (miscarriage and stillbirth). Due to the very small number of stillbirths, the latter will be considered together with miscarriages. Live birth will then be analyzed with logistic regression. The other binary outcomes (preterm birth, low birth weight) will also be analyzed with logistic regression.

In case of numerical issues due to a too small number of events and/or too high number of covariates, we may consider simpler models with less covariates. The models will contain at least the exposure and vaginal dysbiosis.

###### 3.2.5 Handling of missing values

We will report the percentage of missing data for all variables. Depending on the amount of missingness for outcome, exposure, and confounders, we will use multiple imputation by chained equations [10] (with the `mi impute chained` command in Stata or the `mice` package in R). This procedure fills in missing values in multiple variables iteratively based on a sequence of univariate imputation methods with fully conditional specification of prediction equations. Predictive mean matching with five neighbors (`pmm, knn(5)`) will be used for continuous, logistic regression (`logit`) for binary and multinomial regression (`mlogit`) for categorical variables.

In particular, the outcome birth (`outcome_b1`) and gestational age (`outcome_GA`) will be considered as missing in case of a termination. Their values will then be imputed.

Based on such chained equations, a total of 50 multiple imputations will be calculated. The regression models built on those 50 imputed data sets will be analyzed using Rubin's rules [11] (with prefix `mi estimate` in Stata).

###### 3.2.6 Handling of women with more than one pregnancy during the enrolment period

In case of multiple pregnancies during the enrolment period, we will use mixed models to consider the within-participant correlations. This will be the main analysis if more than 5% of participating women have multiple pregnancies.

###### 3.2.7 Sensitivity analyses

In the analysis of categorical gestational age with ordinal or multinomial logistic regression (see paragraph 3.2.3), we will report a marginal effect for each category, by distinguishing between women with vaginal dysbiosis and women without.

Another sensitivity analysis will consist in replacing the presence of *C. trachomatis, N. gonorrhoeae,* and *T. vaginalis* by the exposure to azithromycin.

##### 3.3 Statistical software

Analyses will be performed with Stata 18.0 (or more recent) or with R version 4.2.2 (or more recent). AI-based tools, such as GitHub Copilot, may be used to support code development. Reports will be generated via R 4.2.2 (or more recent), Sweave and LaTeX.

### 5. Annex

Figure 1. Distribution of gestational age at birth, based on 537 observations

A. Histogram

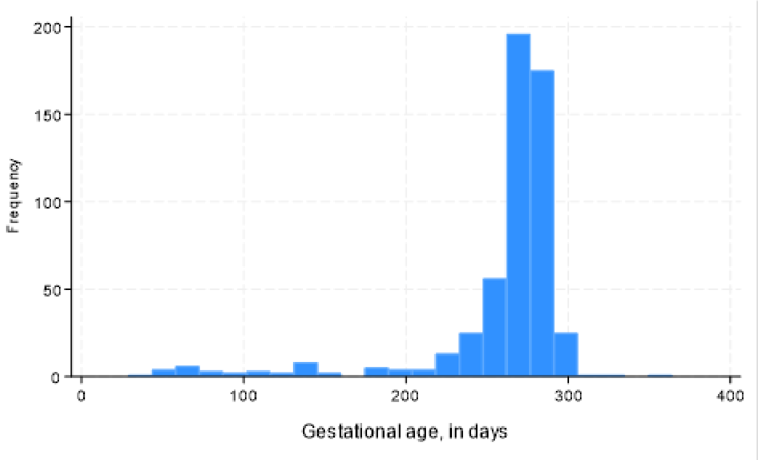

B. Box plot, with median

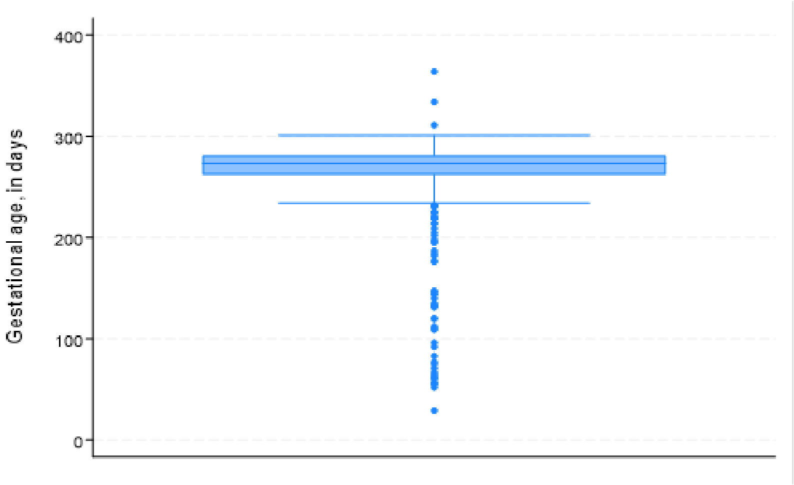

| **CTU Bern** | SAP for: InfPreg | Version: 3.0 |
|---|---|---|
