## supplementary material, file 3 for "*Mycoplasma genitalium* infection and adverse pregnancy outcomes among pregnant women in South Africa: prospective cohort study"

### Supplementary file 3: supplementary tables and figures

***Table S1:*** **Unadjusted analysis of gestational age at birth, according to baseline characteristics**

| **Characteristic** | **Gestational age at birth, in weeks+days** | | | **p value** |
|---|---|---|---|---|
| | **Median** | **IQR** | **Min, max** | |
| All | 39+0 | 37+4, 40+1 | 4+1, 44+3 | |
| Age | | | | 0.01 |
| Younger than 25 years | 39+3 | 38+0, 40+3 | 8+5, 43+0 | |
| 25 to 35 years old | 39+0 | 37+5, 40+1 | 4+1, 43+6 | |
| 35 years or older | 38+5 | 36+2, 39+6 | 7+3, 44+3 | |
| Educational level | | | | 0.83 |
| Less than grade 10 | 39+2 | 38+2, 40+0 | 8+5, 43+6 | |
| Grade 10 or 11 | 39+0 | 37+6, 40+1 | 7+3, 42+6 | |
| Grade 12 or higher | 39+1 | 37+2, 40+2 | 4+1, 44+3 | |
| Alcohol consumption | | | | 0.95 |
| No | 39+0 | 37+3, 40+1 | 4+1, 44+3 | |
| Yes | 39+0 | 38+0, 40+1 | 7+6, 43+6 | |
| Living with HIV | | | | 0.40 |
| No | 39+0 | 37+4, 40+1 | 7+3, 43+6 | |
| Yes, on ART | 38+6 | 37+2, 40+1 | 4+1, 41+6 | |
| Yes, not on ART | 39+3 | 38+3, 40+3 | 17+1, 44+3 | |
| Syphilis | | | | 0.83 |
| No | 39+0 | 37+4, 40+1 | 4+1, 44+3 | |
| Yes | 39+0 | 37+1, 40+4 | 8+1, 42+0 | |
| At least one prior preterm birth | | | | 0.42 |
| No | 39+0 | 37+4, 40+1 | 4+1, 43+6 | |
| Yes | 38+5 | 37+2, 39+5 | 21+0, 44+3 | |
| Vaginal dysbiosis | | | | 0.55 |
| Nugent 0-3 | 39+0 | 37+2, 40+1 | 7+3, 44+3 | |
| Nugent 7-10 | 39+0 | 37+5, 40+2 | 4+1, 43+0 | |
| *C. trachomatis* | | | | 0.13 |
| Negative | 39+0 | 37+3, 40+1 | 4+1, 44+3 | |
| Positive | 39+2 | 38+3, 40+2 | 7+6, 43+6 | |
| *N. gonorrhoeae* | | | | 0.15 |
| Negative | 39+0 | 37+4, 40+1 | 4+1, 44+3 | |
| Positive | 38+5 | 36+2, 39+4 | 13+5, 40+4 | |
| *T. vaginalis* | | | | 0.23 |
| Negative | 39+0 | 37+4, 40+2 | 7+3, 44+3 | |
| Positive | 38+5 | 37+3, 39+6 | 4+1, 43+0 | |
| *Candida* spp. | | | | 0.39 |
| Negative | 39+1 | 37+4, 40+2 | 4+1, 44+3 | |
| Positive | 38+6 | 37+4, 40+0 | 8+0, 42+2 | |
| Azithromycin treatment for *C. trachomatis* | | | | 0.96 |
| No | 39+0 | 37+3, 40+1 | 4+1, 44+3 | |
| Yes | 39+0 | 37+6, 40+1 | 7+6, 43+0 | |

Data are median and interquartile range (IQR); Min=minimum; Max=maximum.

***Table S2: M. genitalium* exposure at 30-34 week follow-up visit and associations with gestational age at birth and other secondary outcomes**

| **Birth outcome** | **Total (N = 441[a])** | ***M. genitalium* negative (N = 415)** | ***M. genitalium* positive (N = 19)** | ***M. genitalium* missing (N = 7)** | **p value** |
|---|---|---|---|---|---|
| Gestational age, in days | | | | | 0.7 |
| Median | 39+2 | 39+2 | 39+4 | 37+5 | |
| Q1, Q3 | 38+1, 40+2 | 38+1, 40+2 | 38+3, 40+0 | 33+1, 39+2 | |
| Min, max | 26+2, 43+0 | 30+4, 43+0 | 37+1, 42+2 | 26+2, 41+4 | |
| Missing, n (%) | 4 | 4 | 0 | 0 | |
| Live birth | | | | | 1 |
| Yes, n (%) | 434 (99%) | 408 (99%) | 19 (100%) | 7 (100%) | |
| No, n (%) | 3 (1%) | 3 (1%) | 0 (0%) | 0 (0%) | |
| Missing, n | 4 | 4 | 0 | 0 | |
| Preterm birth | | | | | 0.09 |
| No, n (%) | 377 (86%) | 354 (86%) | 19 (100%) | 4 (57%) | |
| Yes, n (%) | 60 (14%) | 57 (14%) | 0 (0%) | 3 (43%) | |
| Missing, n | 4 | 4 | 0 | 0 | 0.85 |
| Birth weight, in grams | | | | | |
| Median | 3144 | 3160 | 3200 | 2760 | |
| Q1, Q3 | 2850, 3500 | 2850, 3505 | 2850, 3475 | 2165, 3035 | |
| Min, max | 1080, 4900 | 1270, 4900 | 2200, 4110 | 1080, 3600 | |
| Missing, n (%) | 20 | 20 | 0 | 0 | |
| Low birth weight | | | | | 0.72 |
| No, n (%) | 368 (87%) | 347 (88%) | 16 (84%) | 5 (71%) | |
| Yes, n (%) | 53 (13%) | 48 (12%) | 3 (16%) | 2 (29%) | |
| Missing, n | 20 | 20 | 0 | 0 | |

[a] 441 of 584 women attended their 30-34 week follow-up visit

Q1, first quartile; Q3, third quartile; Min, minimum; Max, maximum.

#### *Table S3* Adjusted estimates of the impact of exposure to *M. genitalium* and vaginal dysbiosis on birth outcomes in all women

| **Outcome** | **Comparison**[a] | **Nugent score 0-3** | **Nugent score 4-10** |
|---|---|---|---|
| Gestational age, in weeks+days | *M. genitalium* not detected | 38+5 (38+2, 39+2) | 39+0 (38+4, 39+3) |
| | *M. genitalium* detected | 39+5 (38+5, 40+4) | 38+3 (37+6, 39+0) |
| | Median difference | 0+6 (-0+2, 2+0) | -0+4 (-1+2, 0+1) |
| Miscarriage or stillbirth | Marginal odds ratio | 1.74 (0.39, 7.72) | 1.74 (0.39, 7.72) |
| | Risk difference | 0.02 (-0.04, 0.08) | 0.02 (-0.05, 0.09) |
| Preterm birth | Marginal odds ratio | 1.04 (0.35, 3.12) | 1.04 (0.35, 3.12) |
| | Risk difference | 0.01 (-0.16, 0.17) | 0.01 (-0.14, 0.15) |
| Birth weight, in grams | *M. genitalium* not detected | 3018 (2821, 3215) | 3099 (2912, 3286) |
| | *M. genitalium* detected | 3116 (2610, 3623) | 3002 (2725, 3279) |
| | Median difference | 98 (-543, 739) | -97 (-430, 236) |
| Low birth weight | Marginal odds ratio | 0.95 (0.25, 3.59) | 0.95 (0.25, 3.59) |
| | Risk difference | -0.01 (-0.19, 0.18) | -0.01 (-0.15, 0.14) |

[a] For gestational age, comparisons are marginal medians and median differences. For other birth outcomes, comparisons are marginal odds ratios and risk differences. Estimates are adjusted for: age, education, alcohol consumption at baseline, previous preterm births, living with HIV, receipt of azithromycin, and exposure to *C. trachomatis, T. vaginalis, N. gonorrhoeae* or *Candida spp*. Quantile regression models include an interaction term between *M. genitalium* and vaginal dysbiosis.

*Table S4:* **Adjusted estimates of the impact of exposure to *M. genitalium* and vaginal dysbiosis on birth outcomes in women with live births.**

| **Outcome**[a] | **Comparison**[b] | **Nugent score 0-3** | **Nugent score 4-10** |
|---|---|---|---|
| Gestational age, in weeks+days | *M. genitalium* not detected | 38+5 (38+1, 39+2) | 39+0 (38+4, 39+3) |
| | *M. genitalium* detected | 39+4 (39+0, 40+1) | 38+4 (38+0, 39+1) |
| | Median difference | 0+6 (0+1, 1+3) | -0+3 (-1+1, 0+1) |
| Preterm birth | Marginal odds ratio | 0.75 (0.18, 3.23) | 0.75 (0.18, 3.23) |
| | Risk difference | -0.04 (-0.21, 0.14) | -0.03 (-0.17, 0.11) |
| Birth weight, in grams | *M. genitalium* not detected | 3010 (2818, 3203) | 3093 (2910, 3275) |
| | *M. genitalium* detected | 3110 (2626, 3594) | 2993 (2710, 3277) |
| | Median difference | 100 (-512, 711) | -99 (-442, 244) |
| Low birth weight | Marginal odds ratio | 0.98 (0.26, 3.72) | 0.98 (0.26, 3.72) |
| | Risk difference | 0 (-0.2, 0.19) | 0 (-0.16, 0.15) |

[a] Estimates are not shown for miscarriage or stillbirth, as this analysis excluded women with these outcomes.

[b] For gestational age, comparisons are marginal medians and median differences. For other birth outcomes, comparisons are marginal odds ratios and risk differences. Estimates are adjusted for: age, education, alcohol consumption at baseline, previous preterm births, living with HIV, receipt of azithromycin, and positive test results for *C. trachomatis*, *T. vaginalis, N. gonorrhoeae* or *Candida spp*. Quantile regression models include an interaction term between *M. genitalium* and vaginal dysbiosis.

***Table S5:*** **Adjusted estimates of the impact of exposure to *M. genitalium* at 30-34 weeks gestation on birth outcomes in women who attended their 30-34 week follow-up visit**

| **Outcome**[a] | **Comparison**[b] | **Estimate** |
|---|---|---|
| Gestational age, in weeks+days | *M. genitalium* not detected | 39+1 (38+5, 39+5) |
| | *M. genitalium* detected | 39+2 (38+4, 40+0) |
| | Median difference | 0+1 (-0+4, 0+6) |
| Birth weight, in grams | *M. genitalium* not detected | 3135.51 (2874.98, 3396.03) |
| | *M. genitalium* detected | 2965.33 (2536.34, 3394.31) |
| | Median difference | -170.18 (-542.65, 202.29) |
| Low birth weight | Marginal odds ratio | 1.57 (0.41, 6.03) |
| | Risk difference | 0 (-0.02, 0.02) |

[a] 143 women who did not attend their 30-34 week follow-up visit are excluded. Since most occurrences of miscarriage, stillbirth or preterm birth occurred before this visit, we do not show estimates for these outcomes and do not present results among women with live births. Analyses are performed on complete cases, excluding women with missing data.

[b] For gestational age, comparisons are marginal medians and median differences. For low birthweight, comparisons are marginal odds ratios and risk differences. Estimates are adjusted for: age, education, alcohol consumption at baseline, previous preterm births, living with HIV, receipt of azithromycin, and exposure to vaginal dysbiosis, *C. trachomatis*, *T. vaginalis*, *N. gonorrhoeae* or *Candida spp.* at follow-up.

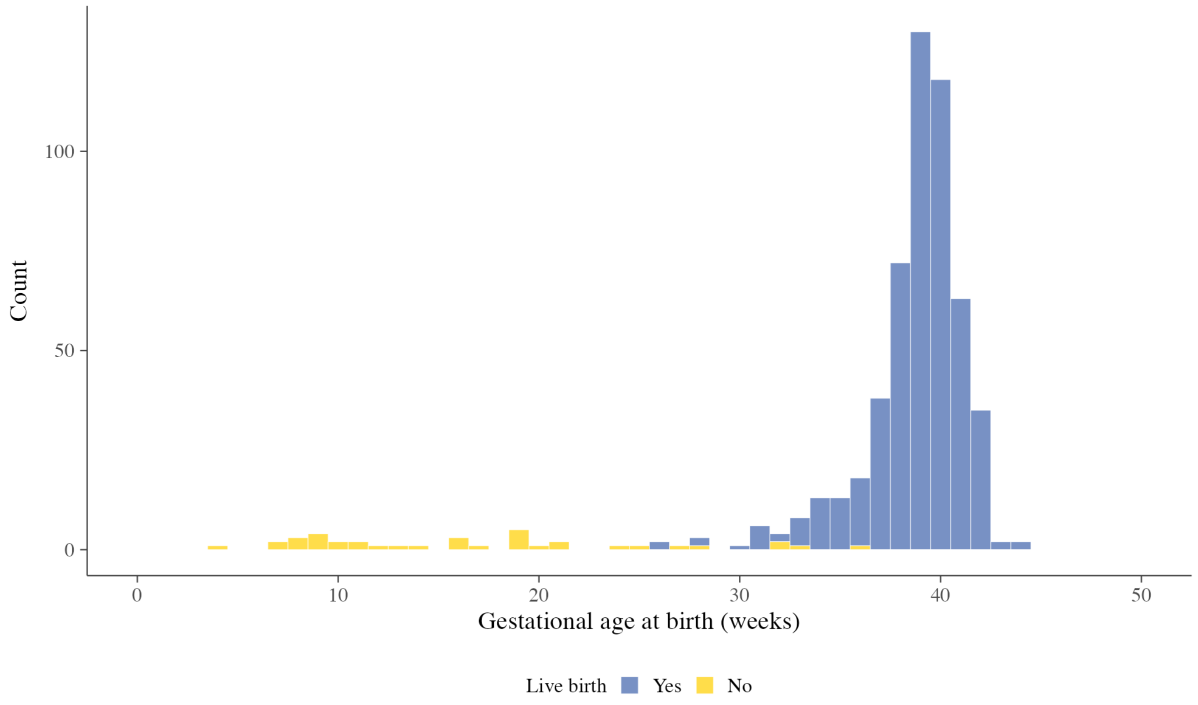

#### *Figure S1*: Histogram of gestational age at delivery for 584 women

Blue bars indicate women with live births, yellow indicates stillbirth, miscarriage or neonatal death

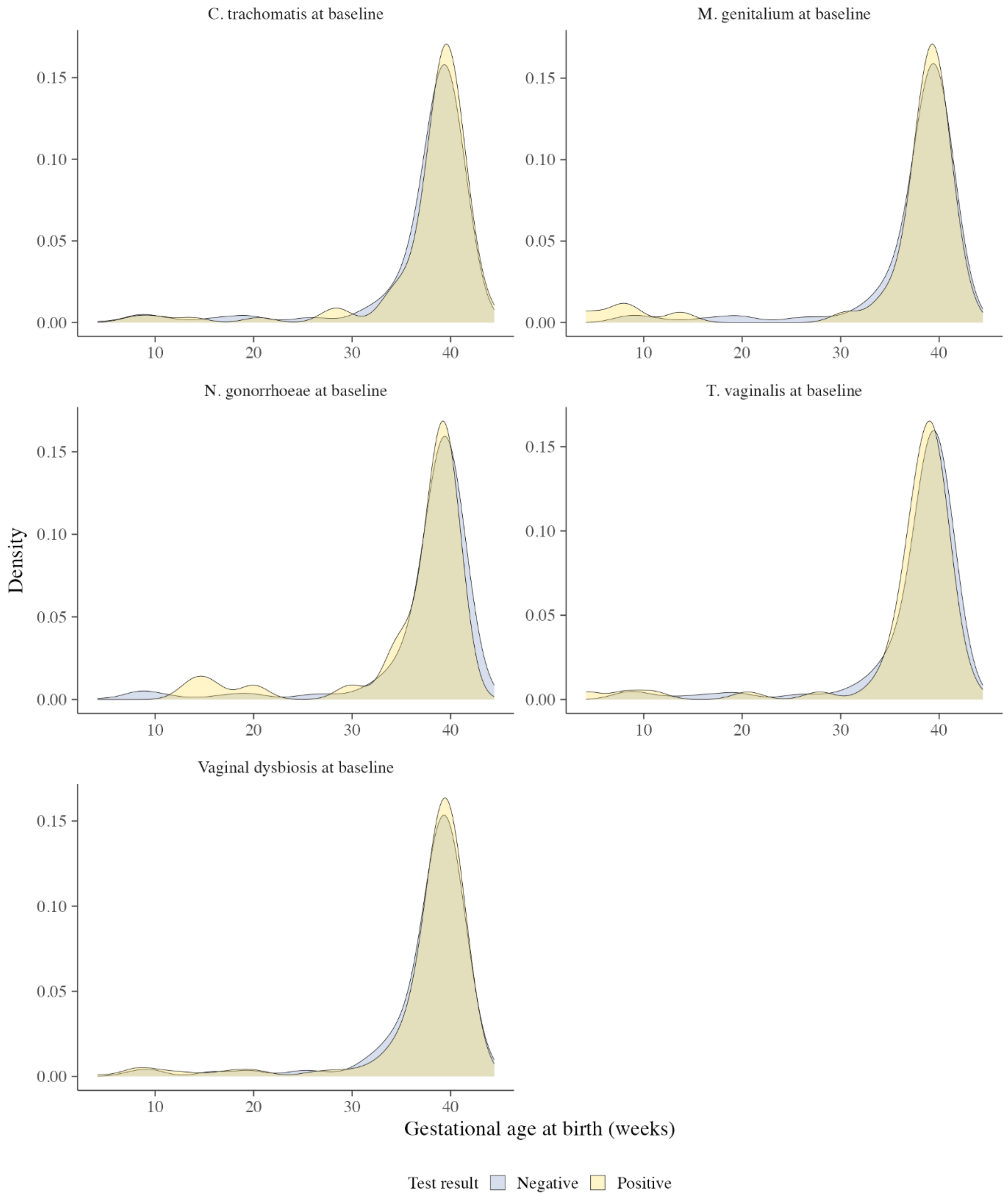

***Figure S2*: Density plots of gestational age at delivery for 560 women, presented separately for each sexually transmitted infection or vaginal dysbiosis**

Blue indicates densities for women who tested negative to the infection, yellow indicates densities for women who tested positive. Densities were estimated with a Gaussian kernel and bandwidth of 1.5.

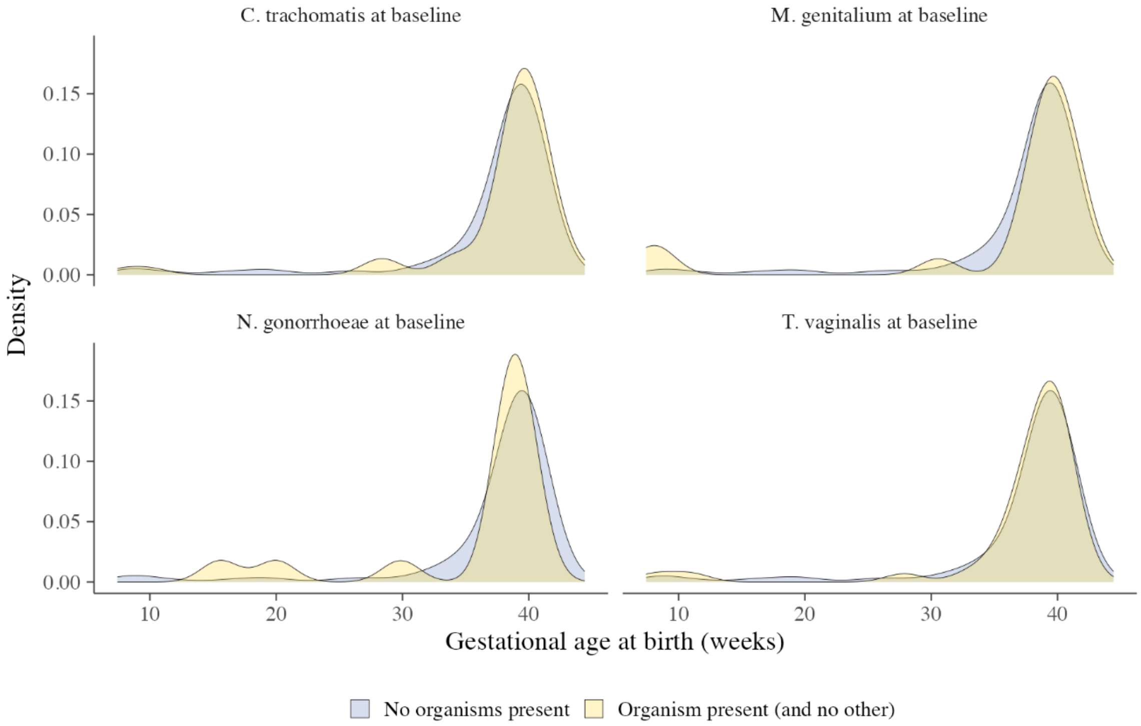

| Count | *C. trachomatis* | *M. genitalium* | *N. gonorrhoeae* | *T. vaginalis* |
|---|---|---|---|---|
| No organisms present | 466 | 504 | 509 | 485 |
| Organism present (and no other) | 58 | 20 | 15 | 39 |

***Figure S3*: Density plots of gestational age at delivery for 524 women who tested positive to each STI (and no other) in comparison to a reference group with no STIs present**

The reference group with no organisms present, shown in blue, includes women who tested negative for all of the following organisms at baseline: *M. genitalium, T. vaginalis, C. trachomatis, N. gonorrhoeae*. The comparison group, shown in yellow, includes women who tested positive at baseline for only the given STI, and no other. Women with missing test results or gestational age at delivery are excluded. Densities were estimated with a Gaussian kernel and bandwidth of 1.5.

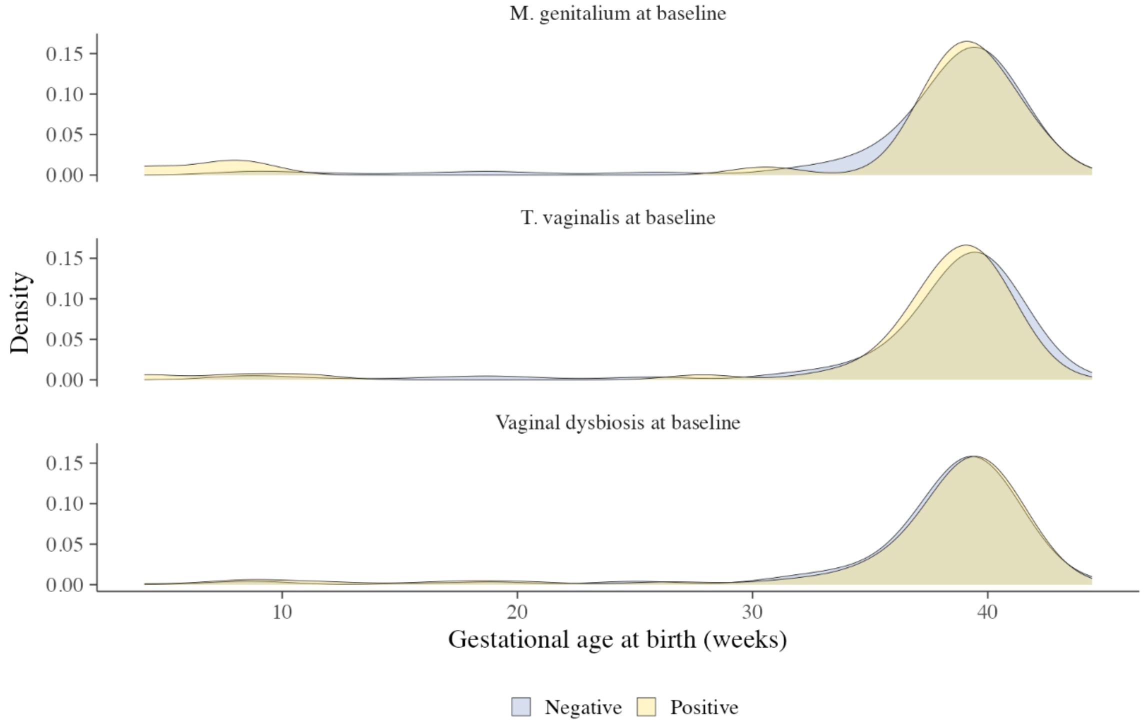

| | *M. genitalium* | *T. vaginalis* | Vaginal dysbiosis |
|---|---|---|---|
| Organism not present | 413 | 396 | 169 |
| Organism present | 27 | 44 | 271 |

***Figure S4*: Density plots of gestational age at delivery for women who did not receive treatment with azithromycin**

Women with missing test results or gestational age at birth are excluded, along with women who received treatment with azithromycin. Density plots for *C. trachomatis* and *N. gonorrhoeae* are not shown due to small counts, as almost all women who tested positive to these two organisms at baseline received treatment. Blue indicates densities for women who tested negative to the infection, yellow indicates densities for women who tested positive. Densities were estimated with a Gaussian kernel and bandwidth of 1.5.

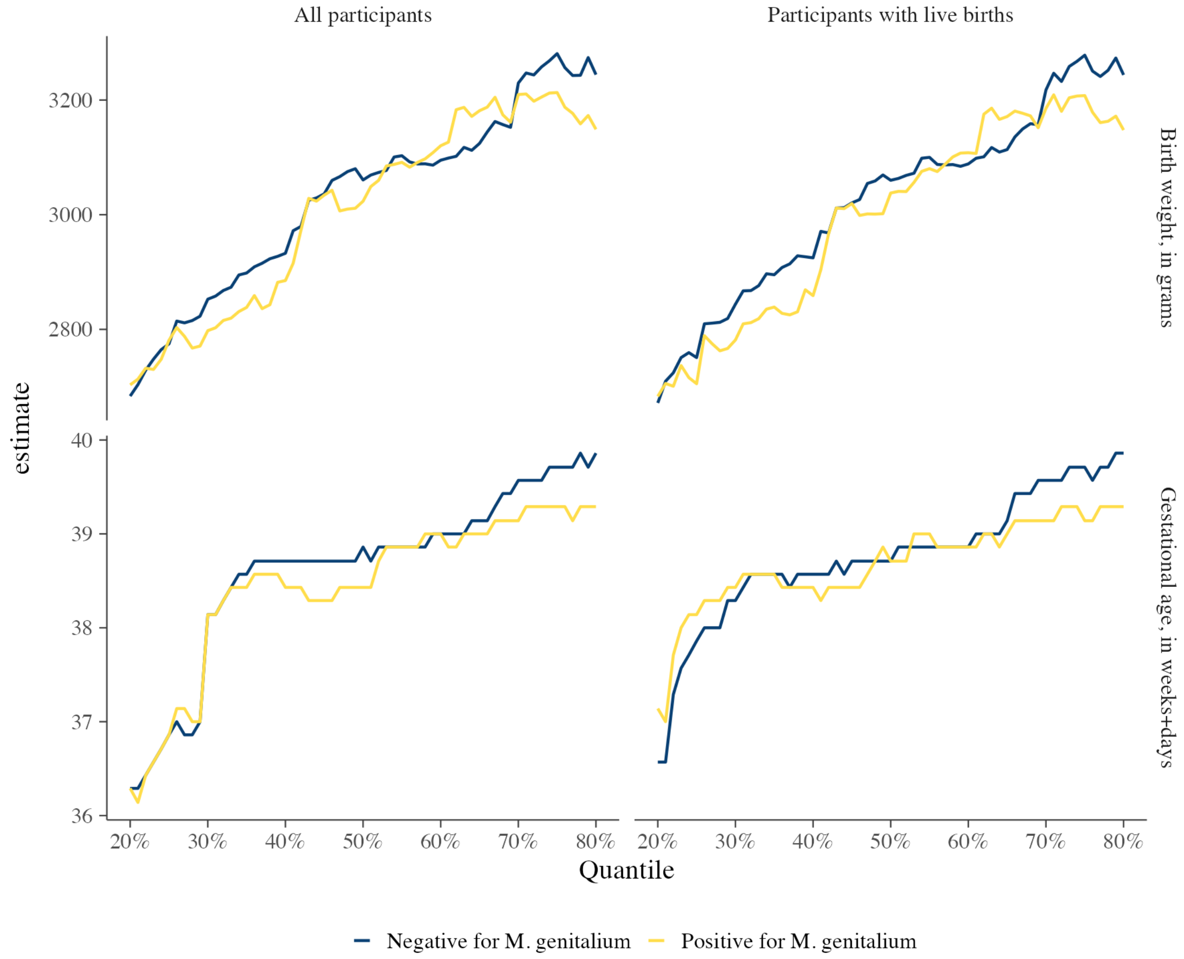

***Figure S5*: Quantile regression estimates for the effect of *M. genitalium* exposure on gestational age at birth, in weeks, and birthweight, in grams**

Lines indicate marginal estimates of gestational age at birth or birth weight for each given quantile, shown separately for women with and without *M. genitalium*. Results are shown for regression estimates fitted to all women (left) and only to women with live births (right).
