## supplementary material, file 1 for "*Mycoplasma genitalium* infection and adverse pregnancy outcomes among pregnant women in South Africa: prospective cohort study"

### Supplementary material 1: STROBE checklist for cohort studies

| Item | | Item | Recommendation | Page, section |
|---|---|---|---|---|
| Title and abstract | | 1 | (a) Indicate the study's design with a commonly used term in the title or the abstract | 1 |
| | | | (b) Provide in the abstract an informative and balanced summary of what was done and what was found | 2 |
| Introduction | | | | |
| | Background/rationale | 2 | Explain the scientific background and rationale for the investigation being reported | 3, Introduction, para 1-3 |
| | Objectives | 3 | State specific objectives, including any prespecified hypotheses | 3, Introduction, para 3 |
| Methods | | | | |
| | Study design | 4 | Present key elements of study design early in the paper | 3-4, Sudy design |
| | Setting | 5 | Describe the setting, locations, and relevant dates, including periods of recruitment, exposure, follow-up, and data collection | 4, Study setting and population |
| | Participants | 6 | (a) Give the eligibility criteria, and the sources and methods of selection of participants. Describe methods of follow-up | 4, Study setting and population; p5, Study procedures and visits |
| | | | (b) For matched studies, give matching criteria and number of exposed and unexposed | Not applicable |
| | Variables | 7 | Clearly define all outcomes, exposures, predictors, potential confounders, and effect modifiers. Give diagnostic criteria, if applicable | 4, Study procedures and visits; 5, Microbiological analyses; 5, Outcomes and statistical analyses |
| | Data sources/measurement | 8 | For each variable of interest, give sources of data and details of methods of assessment (measurement). Describe comparability of assessment methods if there is more than one group | 4, Study procedures and visits; 5, Microbiological analyses; 5, Outcomes and statistical analyses |
| | Bias | 9 | Describe any efforts to address potential sources of bias | 3, Methods, 6, Outcomes and statistical analyses |
| | Study size | 10 | Explain how the study size was arrived at | 5, Outcomes and statistical analyses |
| | Quantitative variables | 11 | Explain how quantitative variables were handled in the analyses. If applicable, describe which groupings were chosen and why | 5-6, Outcomes and statistical analyses |
| | Statistical methods | 12 | (a) Describe all statistical methods, including those used to control for confounding | 6, Outcomes and statistical analyses |
| | | | (b) Describe any methods used to examine subgroups and interactions | 6, Outcomes and statistical analyses |
| | | | (c) Explain how missing data were addressed | 6, Outcomes and statistical analyses |
| | | | (d) If applicable, explain how loss to follow-up was addressed | 6, Outcomes and statistical analyses |
| | | | (e) Describe any sensitivity analyses | 6, Outcomes and statistical analyses |
| Results | | | | |
| | Participants | 13 | (a) Report numbers of individuals at each stage of study—eg numbers potentially eligible, examined for eligibility, confirmed eligible, included in the study, completing follow-up, and analysed | 6, Results |
| | | | (b) Give reasons for non-participation at each stage | 7, Results |
| | | | (c) Consider use of a flow diagram | 7, Results |
| | Descriptive data | 14 | (a) Give characteristics of study participants (eg demographic, clinical, social) and information on exposures and potential confounders | Table 1 |
| | | | (b) Indicate number of participants with missing data for each variable of interest | Table 1 |
| | | | (c) Summarise follow-up time (eg, average and total amount) | 7, Reported totals followed up to birth |
| | Outcome data | 15 | Report numbers of outcome events or summary measures over time | Table 2, Table S1 |
| | Main results | 16 | (a) Give unadjusted estimates and, if applicable, confounder-adjusted estimates and their precision (eg, 95% confidence | 9-11, Tables 2 and 3, Table S1-S5, Fig S2-S4 |

| Item | | Item | Recommendation | Page, section |
|---|---|---|---|---|
| | | | interval). Make clear which confounders were adjusted for and why they were included | |
| | | | (b) Report category boundaries when continuous variables were categorized | Table 2 |
| | | | (c) If relevant, consider translating estimates of relative risk into absolute risk for a meaningful time period | Not applicable |
| | Other analyses | 17 | Report other analyses done—eg analyses of subgroups and interactions, and sensitivity analyses | 9, 12 |
| Discussion | | | | |
| | Key results | 18 | Summarise key results with reference to study objectives | 12 |
| | Limitations | 19 | Discuss limitations of the study, taking into account sources of potential bias or imprecision. Discuss both direction and magnitude of any potential bias | 13 |
| | Interpretation | 20 | Give a cautious overall interpretation of results considering objectives, limitations, multiplicity of analyses, results from similar studies, and other relevant evidence | 13-14 |
| | Generalisability | 21 | Discuss the generalisability (external validity) of the study results | 15 |
| Other information | | | | |
| | Funding | 22 | Give the source of funding and the role of the funders for the present study and, if applicable, for the original study on which the present article is based | 16 |

**Source:** https://www.equator-network.org/reporting-guidelines/strobe/
